# Three Dimensions of Compounding Neglect: How Biobanks, Clinical Trials, and Scientific Literature Systematically Exclude the Global South

**DOI:** 10.64898/2026.02.10.26346004

**Authors:** Manuel Corpas, Maxim B. Freidin, Julio Valdivia-Silva, Simeon Baker, Segun Fatumo, Heinner Guio

## Abstract

Global health inequities are widely documented in outcomes. However, the research systems that generate knowledge, trials, and discovery have rarely been evaluated as an integrated structure. We introduce the Health Equity Informative Metrics (HEIM) framework, a three-dimensional audit of discovery (biobank output), translation (clinical trial activity), and knowledge (semantic organisation of the scientific literature). Analysing 70 international biobanks, 563,725 registered clinical trials, 13.1 million PubMed abstracts, and 175 Global Burden of Disease categories, we demonstrate that exclusion compounds systematically for diseases that primarily burden the Global South. No WHO-classified neglected tropical disease has generated a publication from these 70 biobanks. Clinical trial sites concentrate 2.5-fold in high-income countries relative to disease burden. Diseases disproportionately affecting low-and middle-income regions are 44% more semantically isolated from mainstream biomedical research than other conditions (P < 0.0001, Cohen’s d = 1.80), limiting cross-disciplinary integration. Nine of the ten most neglected diseases across all dimensions disproportionately affect the Global South, and these disparities show no improvement over 26 years. By contrast, the trajectory of HIV/AIDS demonstrates that sustained, coordinated investment can reverse semantic isolation and integrate a once-marginalised disease into mainstream biomedical networks. HEIM reframes research inequity as a measurable, multi-stage enterprise and establishes a framework for health data accountability.

## Introduction

Lymphatic filariasis affects 120 million people worldwide ^1^, causing debilitating swelling that traps individuals in cycles of poverty and stigma. Yet across 70 major international biobanks ^2^, we found no publications, genomic studies or translational research pipelines for this debilitating disease. We contend this is not an isolated failure, and that it reflects a systematic pattern of exclusion embedded in the architecture of biomedical research itself.

While more than 80% of premature deaths from noncommunicable diseases occur in low-and middle-income countries ^3^, the biomedical research enterprise remains overwhelmingly oriented toward conditions prevalent in wealthy nations ^4–6^. This persistent structural exclusion, where the diseases of poverty remain invisible to the research infrastructure that could address them ^7,8^, has continued for decades despite sustained advocacy and targeted funding initiatives ^9,10^. Health outcome inequities are extensively documented ^11–13^, but the structural biases embedded within research infrastructure itself remain poorly understood and inadequately measured ^14,15^. Recent work has quantified the mismatch between publication output and disease burden at scale ^5^, yet single-dimension analyses cannot reveal whether disadvantage compounds across the research pipeline. Critically, the HIV/AIDS experience demonstrates that sustained investment can fundamentally reverse structural marginalisation ^16^: a disease once neglected from mainstream biomedicine is now deeply integrated into immunology, virology, and global health ^17–20^.

Research inequity operates at multiple stages of the scientific enterprise ^21,22^. At the discovery stage, biobanks and cohort studies generate foundational data for genomic medicine ^23–25^, yet concentrate overwhelmingly in high-income countries and European-ancestry populations ^26–28^. At the translation stage, trial sites cluster in wealthy nations regardless of where disease burden is greatest ^29–31^. At the knowledge stage, bibliometric and semantic methods exist but have rarely assessed whether specific diseases occupy marginalised positions within published science ^32,33^. Previous single-dimension metrics, including publication counts ^34^, trial registrations ^29^, and funding allocation ^35^, cannot capture compounding disadvantage, whereby diseases neglected in early-stage research encounter systematic barriers in downstream development and implementation ^36–38^.

Here we introduce the Health Equity Informative Metrics (HEIM) framework (**Fig. 1**), a three-dimensional analysis of biomedical research equity encompassing **discovery** (biobank research output), **translation** (clinical trial data), and **knowledge** (semantic structure of scientific literature). We analysed 70 biobanks from the International Health Cohorts Consortium across 29 countries ^2^, 563,725 registered clinical trials ^39^, and 13.1 million PubMed abstracts spanning 26 years (2000-2025). Using the Global Burden of Disease 2021 taxonomy to classify 175 disease categories ^40^, we harmonised all three data sources to measure the compounding structure of research neglect across the biomedical pipeline.

**Figure 1.**
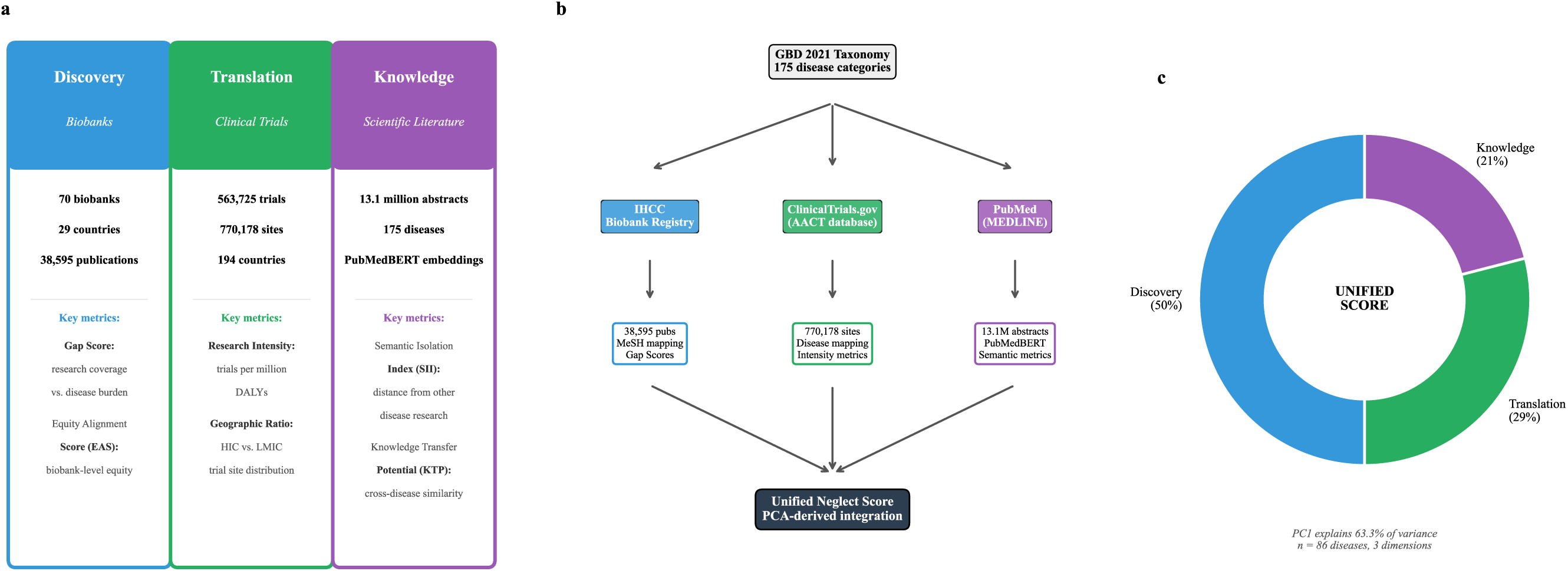
The HEIM framework integrates three dimensions of research equity. Schematic overview of the Health Equity Informative Metrics (HEIM) framework. (a) Three dimensions of research equity with data scale and key metrics for each: **Discovery** measures biobank research coverage relative to disease burden (*Gap Score*) and biobank-level equity (*Equity Alignment Score, EAS*); **Translation** quantifies clinical trial activity per unit of disease burden (Research Intensity) and the distribution of trial sites between high-income and low-and middle-income countries (*Geographic Ratio*); **Knowledge** captures how disconnected a disease’s research is from the broader biomedical literature (*Semantic Isolation Index, SII*) and the potential for cross-disease methodological transfer (*Knowledge Transfer Potential, KTP*). (b) Data pipeline from the GBD 2021 taxonomy through three primary sources (IHCC Biobank Registry, ClinicalTrials.gov via the AACT database, and PubMed/MEDLINE) to dimension-specific metrics. (c) Integration into a *Unified Neglect Score* using empirically derived weights from principal component analysis (PCA): Discovery = 0.50, Translation = 0.29, Knowledge = 0.21 (PC1 explains 63.3% of variance across 86 diseases with complete three-dimensional data).

## Results

### The Discovery Dimension: Biobank Research Shows Severe Equity Gaps

Only 1 of 70 major international biobanks achieves high equity alignment with global disease burden. This finding emerged from our analysis of 70 biobanks registered with the International Health Cohorts Consortium (IHCC), spanning 29 countries across six WHO regions (**Fig. 2a**). These cohorts have collectively generated 38,595 Global Burden of Disease-specific publications indexed in PubMed (Methods). To quantify alignment between research output and global disease burden, we developed the Equity Alignment Score (EAS), which penalises biobanks for critical gaps in high-burden diseases while accounting for research capacity (Methods).

**Figure 2.**
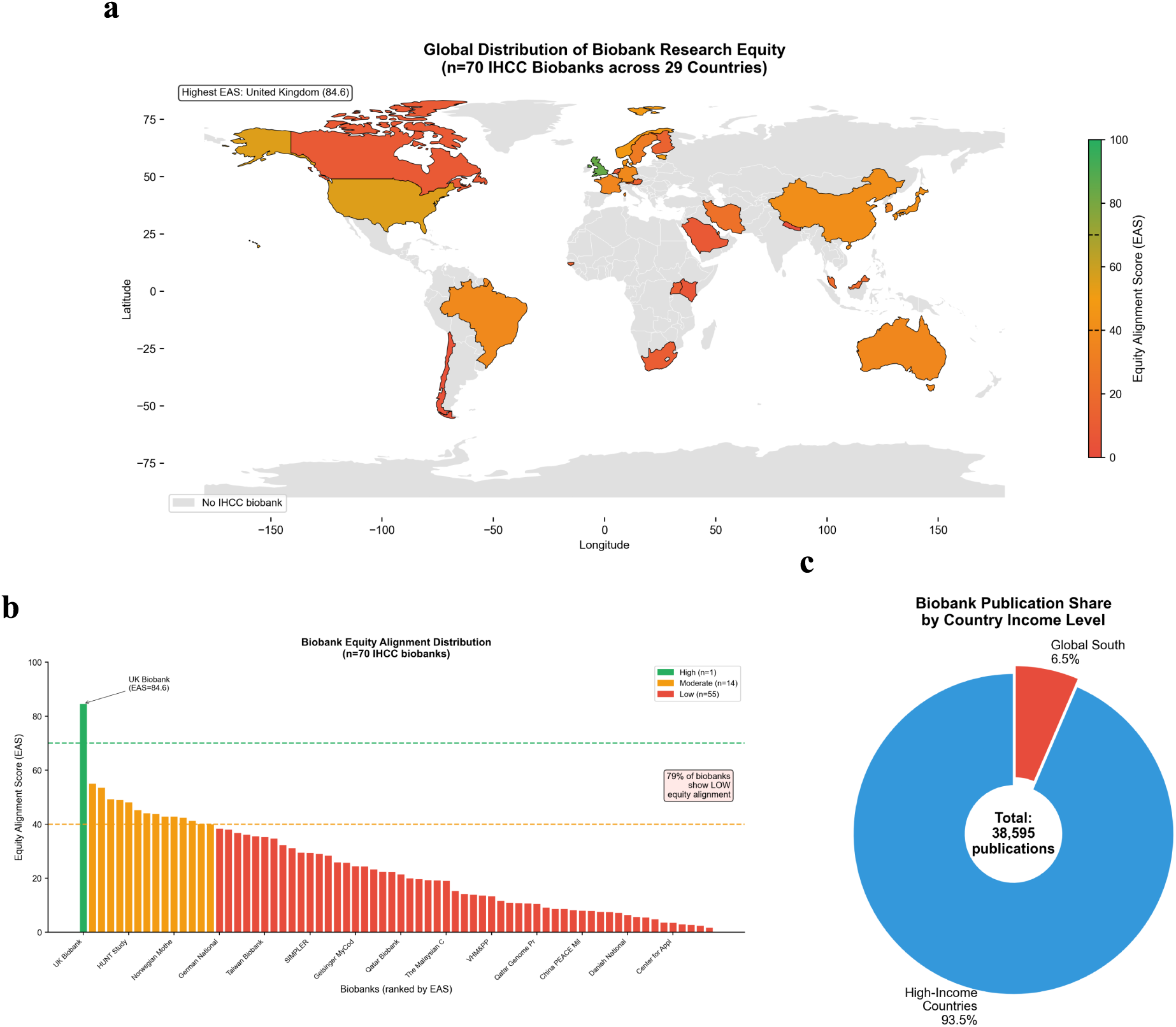
Only one of 70 global biobanks achieves equitable disease coverage. The Equity Alignment Score (EAS) measures how well a biobank’s disease-specific publication output matches global disease burden, combining gap severity, burden coverage, and capacity (scale 0-100; High ≥70, Moderate 40-69, Low <40). (a) World map showing locations of 70 IHCC biobanks, coloured by EAS category. (b) Distribution of EAS across biobanks. Only UK Biobank (EAS = 84.6) achieves High alignment; 55 of 70 biobanks (79%) score Low. (c) Publication share by country income group, showing 93.5% concentration in high-income countries.

The distribution of equity alignment was heavily skewed toward poor performance. Only UK Biobank ^24^ achieved “High” equity alignment (EAS = 84.6), defined as EAS ≥ 70 (**Fig. 2b**). Fourteen biobanks showed “Moderate” alignment (EAS 40-69), including Nurses’ Health Study ^41^ (55.1), Women’s Health Initiative ^42^ (53.5), and Estonian Biobank ^43^ (49.3). The remaining 55 biobanks (79%) demonstrated “Low” alignment (EAS < 40), indicating substantial misalignment between their research portfolios and global health priorities.

Geographic concentration was pronounced. European and North American biobanks accounted for 45 of 70 cohorts (64%) and 33,915 of 38,595 publications (87.9%). The six African biobanks collectively produced 277 publications (0.7%), despite Africa bearing 24% of the global disease burden ^44^: a 34-fold disparity between research output and disease burden. High-income country biobanks generated 93.5% of all publications, while biobanks in Global South countries contributed just 6.5% (**Fig. 2c**; **Extended Data Fig. 3**).

We identified 22 diseases with “Critical” research gaps (Gap Score > 70), corresponding to severe underrepresentation relative to disease burden. These included malaria (17 publications across all biobanks, despite 55.2 million disability-adjusted life years [DALYs] globally), dengue (0 publications, 2.1 million DALYs), schistosomiasis (0 publications, 1.7 million DALYs), and lymphatic filariasis (0 publications, 1.3 million DALYs). In particular, diseases such as malaria and schistosomiasis have substantial absolute research volume in the wider literature; their high Gap Scores reflect not an absence of research but a structural disconnect from major biobank cohorts, which have not incorporated these conditions into their research portfolios despite decades of dedicated study outside the biobank context. An additional 26 diseases showed “High” gaps (Gap Score 50–70), and 48 showed “Moderate” gaps (Gap Score 30–50). Only 83 of the 179 disease categories analysed in the Discovery dimension (46%) had “Low” gaps indicating adequate coverage relative to burden.

Having established these gaps in discovery-stage research, we next examined whether similar patterns emerge in clinical translation.

### The Translation Dimension: Clinical Trials Concentrate in high-income settings

Analysis of 563,725 clinical trials registered on ClinicalTrials.gov (2000-2025), with 770,178 geolocated trial site records, revealed systematic geographic concentration (**Fig. 3a**). Trial sites in high-income countries numbered 552,952 (71.8%), compared to 217,226 (28.2%) in low-and middle-income countries, representing a 2.5-fold disparity.

**Figure 3.**
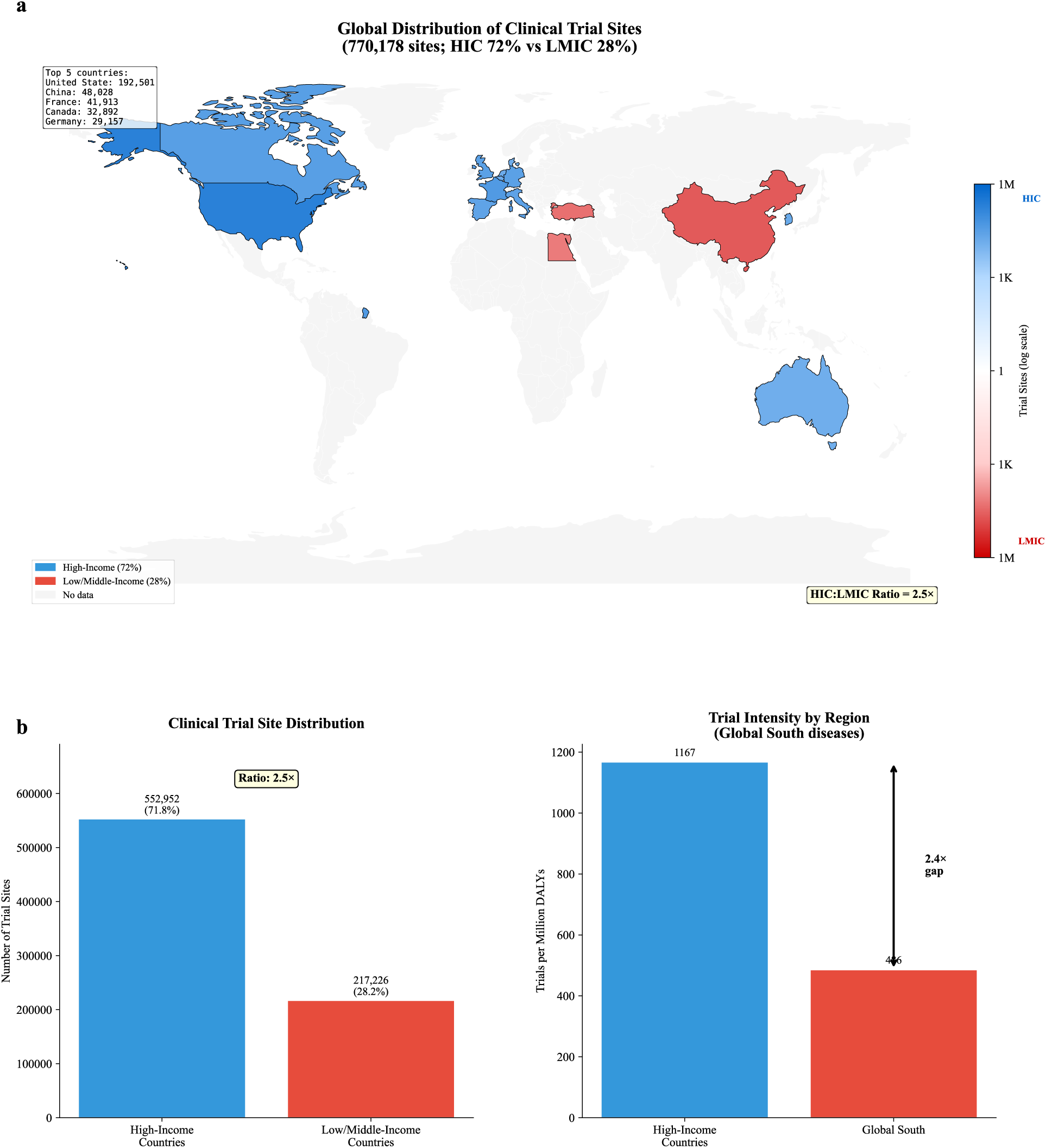
Clinical trial sites concentrate 2.5-fold in high-income countries. (a) Global heatmap of 770,178 clinical trial sites, showing concentration in North America and Europe. (b) Trial intensity comparison between high-income countries and Global South, demonstrating 2.4-fold intensity gap.

The United States alone hosted 192,501 trial sites (25.0% of the global total), followed by China (48,028; 6.2%), France (41,913; 5.4%), and Canada (32,892; 4.3%). The ten countries with the most trial sites were predominantly high-income, with China and Turkey as the only middle-income exceptions in the top fifteen. China’s position reflects its status as an emerging global research power with rapidly expanding clinical trial infrastructure, though this capacity has not translated into proportional representation for neglected diseases affecting other LMIC regions.

We examined 89 disease categories with sufficient trial data for geographic analysis. For the 30 diseases predominantly affecting the Global South (identified through GBD burden distributions) we found a substantial intensity gap. Global South countries hosted 20.3% of clinical trials for these conditions but bear 38.0% of the associated disability-adjusted life years (DALYs), yielding a trial intensity ratio of 485.9 trials per million DALYs compared to 1,167.4 in high-income countries (intensity gap = 2.4-fold; **Fig. 3b**).

Neglected tropical diseases (NTDs) showed the most severe underrepresentation. For lymphatic filariasis (1.3 million DALYs), only 89 trials were registered globally, compared to 47,892 for diabetes mellitus (78.9 million DALYs): a 538-fold disparity. What limited NTD research exists does occur where disease is prevalent (71% of lymphatic filariasis sites in endemic countries), but absolute volumes remain critically insufficient. Schistosomiasis (1.7 million DALYs) had 156 trials; cysticercosis (1.2 million DALYs) had 47 trials. By contrast, ischemic heart disease (188.4 million DALYs) had 38,441 trials, representing approximately 204 trials per million DALYs compared to just 68 for lymphatic filariasis.

NTD-mapped trials account for just 3,703 of 563,725 registered trials (0.66%). The nature of this research reveals a second layer of disadvantage. Of NTD trials, 39.5% are observational, compared to 23.2% for other conditions, consistent with a pipeline that generates epidemiological data but fails to advance treatments. NTD Phase 1 trials are underrepresented (5.2% versus 8.3% for non-NTD conditions), suggesting fewer drug candidates entering clinical testing. Late-phase trials (Phase 3 and 4) constitute 13.2% of NTD trials compared to 13.3% for non-NTD conditions; the apparent parity masks vastly smaller absolute numbers. Praziquantel has remained the sole treatment for schistosomiasis for over four decades, with no Phase III alternatives despite the disease affecting 240 million people. Onchocerciasis, affecting 21 million people, has only 34 registered trials ^45^, with just 23.5% in late-phase: a disease whose therapeutic pipeline has been essentially stalled for decades.

Academic and non-profit institutions lead 75.7% of all clinical trials as primary sponsors, industry leads 22.4%, and the NIH leads only 1.9% (10,691 trials). The concentration of industry sponsorship in commercially viable conditions and the reliance of NTD research on academic and philanthropic funding creates a structural vulnerability: when philanthropic attention shifts, NTD trial activity declines relative to other conditions. Only 13.4% of all trials report participant demographic data, rendering the representativeness of trial populations unmeasurable at scale.

This opacity is itself an equity concern: without demographic reporting, it is impossible to assess whether the trials that do exist for Global South diseases actually enrol the populations most affected. The share of clinical trials addressing Global South priority diseases declined from 43.1% in 2020 (inflated by COVID-19) to 32.5% in 2024, the lowest recorded percentage in a 15-year window.

Beyond research volume and geography, we asked whether neglected diseases also occupy marginalised positions in the structure of scientific knowledge itself.

### The Knowledge Dimension: Neglected Diseases Occupy Isolated Semantic Space

To assess structural marginalisation in the research literature beyond simple publication counts, we generated semantic embeddings for 13.1 million PubMed extended abstracts spanning 175 GBD disease categories (2000-2025). Using PubMedBERT ^46^, we represented each abstract as a 768-dimensional vector capturing its semantic content, then computed disease-level centroids and inter-disease similarity matrices (Methods).

We developed three novel metrics to characterise the semantic landscape (Methods). The **Semantic Isolation Index (SII)** measures the mean cosine distance from a disease’s PubMedBERT centroid to its 100 nearest-neighbour disease centroids; higher values indicate greater structural disconnection from mainstream biomedical discourse. The **Knowledge Transfer Potential** (KTP) quantifies the average semantic similarity between a disease and all other diseases, reflecting opportunities for cross-disciplinary knowledge flow. The **Research Clustering Coefficient** (RCC) measures the mean cosine distance from a disease’s individual abstracts to its centroid; lower values indicate tightly focused research, while higher values suggest a more heterogeneous literature.

UMAP projection of 175 disease centroids in semantic space revealed clear separation of NTDs from mainstream biomedical research clusters (**Fig. 4a**). NTDs formed a distinct cluster removed from cardiovascular, oncological, and neurological disease research, indicating that the language of NTD research is structurally disconnected from the dominant knowledge domains. This pattern was consistent at the category level: NTDs exhibited the highest median SII among all GBD Level 2 disease categories (**Fig. 4b**).

**Figure 4.**
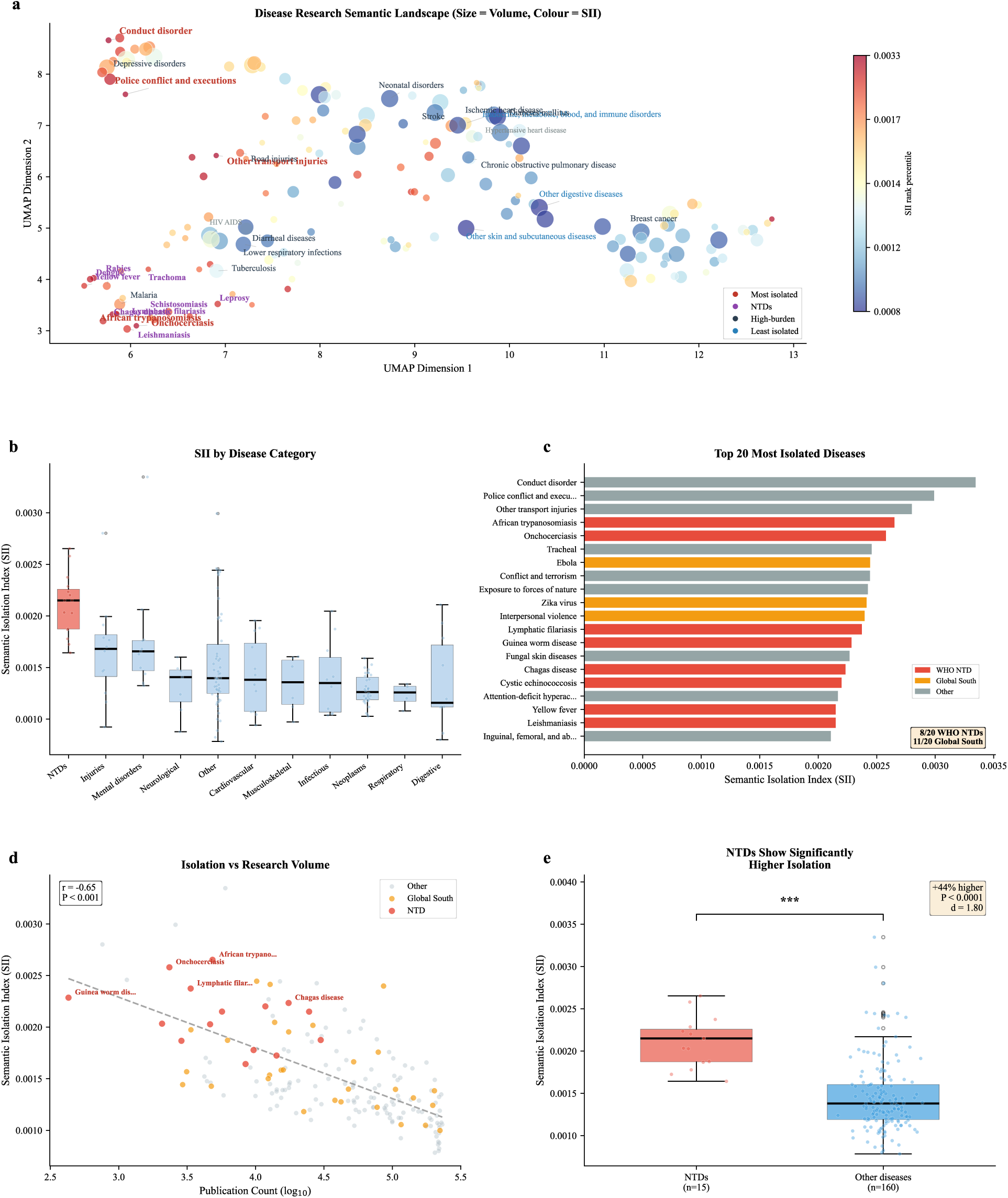
Neglected tropical diseases are semantically isolated from mainstream biomedical research. (a) UMAP projection of 175 diseases in semantic space; point size reflects research volume and colour encodes SII rank percentile, showing NTD cluster separation. (b) Distribution of SII across 11 GBD disease categories; NTDs show the highest median isolation. (c) Top 20 most semantically isolated GBD diseases, coloured by WHO NTD status (red), Global South priority (orange), or other (grey); 8 of the 20 are WHO NTDs and 11 are Global South conditions. (d) Semantic isolation decreases with research volume (Pearson r = −0.65, P < 0.001), but NTDs remain outliers above the regression line. (e) NTDs (n = 15; the 15 of 19 WHO NTDs with sufficient PubMed abstracts for embedding; see Methods) have significantly higher SII than other diseases (n = 160; Welch t-test P < 0.0001, Cohen’s d = 1.80). SII is the mean cosine distance from each disease’s PubMedBERT centroid to its 100 nearest-neighbour centroids; higher values indicate greater semantic isolation.

The top 20 most semantically isolated diseases were dominated by Global South conditions: 8 of 20 are WHO-classified NTDs and 11 are Global South priority diseases (**Fig. 4c**). Semantic isolation was not simply a function of research volume; the negative correlation between volume and SII (Pearson r = −0.65, P < 0.001) showed NTDs as consistent outliers above the regression line (**Fig. 4d**). The NTD versus non-NTD comparison confirmed a 44% elevation in mean SII (0.00211 vs 0.00146; Welch’s t-test, P < 0.0001, Cohen’s d = 1.80; **Fig. 4e**). By contrast, HIV/AIDS ranked among the 25% least isolated diseases despite its origins as a disease of poverty, a pattern we examine further in the Discussion.

Semantic isolation correlated significantly with Discovery dimension Gap Scores (Pearson r = 0.67, P < 0.001), indicating that diseases neglected in biobank research also occupy marginalised positions in the broader scientific literature. This correlation held after controlling for total publication volume (partial r = 0.58, P < 0.001), confirming that isolation reflects qualitative positioning, not merely quantity.

### HIV/AIDS: Empirical Evidence That Semantic Isolation Can Be Reversed

HIV/AIDS provides empirical evidence that a disease of poverty can achieve semantic integration with mainstream biomedicine. Despite originating in marginalised populations, HIV/AIDS now ranks among the 25% least isolated diseases (41st of 175) and shows the highest Knowledge Transfer Potential (KTP = 0.999) of any infectious disease, indicating dense semantic connections to immunology, virology, and clinical trial methodology. This contrasts sharply with NTDs, which remain in the top quartile of isolation despite decades of WHO programmatic attention. The difference suggests that sustained research investment, not merely disease recognition, drives semantic integration.

The semantic integration of HIV/AIDS was not uniform across knowledge domains. HIV research showed the strongest connectivity gains with immunology (KTP increase: 0.034), clinical trial methodology (0.028), and global health systems research (0.025). These are precisely the domains where PEPFAR, the Global Fund, and other coordinated investments concentrated capacity-building efforts ^47^. By contrast, HIV’s connectivity with neglected disease research remained low (KTP: 0.011), suggesting that integration benefits did not diffuse to other diseases of poverty. This pattern indicates that strategic investment can reduce isolation for targeted diseases but does not automatically benefit adjacent neglected conditions.

### Unified Neglect Score: Identifying Compounding Disadvantage

To identify diseases experiencing systematic neglect across all dimensions, we computed a **Unified Neglect Score** integrating Discovery (Gap Score), Translation (clinical trial equity), and Knowledge (Semantic Isolation Index) with empirically derived weights from principal component analysis (see Methods). The score ranges from 0 (no neglect) to 100 (maximum neglect across all dimensions).

We computed Unified Scores for all 175 GBD Level 3 diseases: 86 with complete data across all three dimensions and 89 using a two-dimension fallback (Discovery and Knowledge only; see Methods). The mean Unified Score was 27.7 (SD = 22.2), with a range from 0.5 (breast cancer) to 87.9 (Guinea worm disease). The distribution was right-skewed, with a long tail of highly neglected conditions (**Fig. 5**).

**Figure 5.**
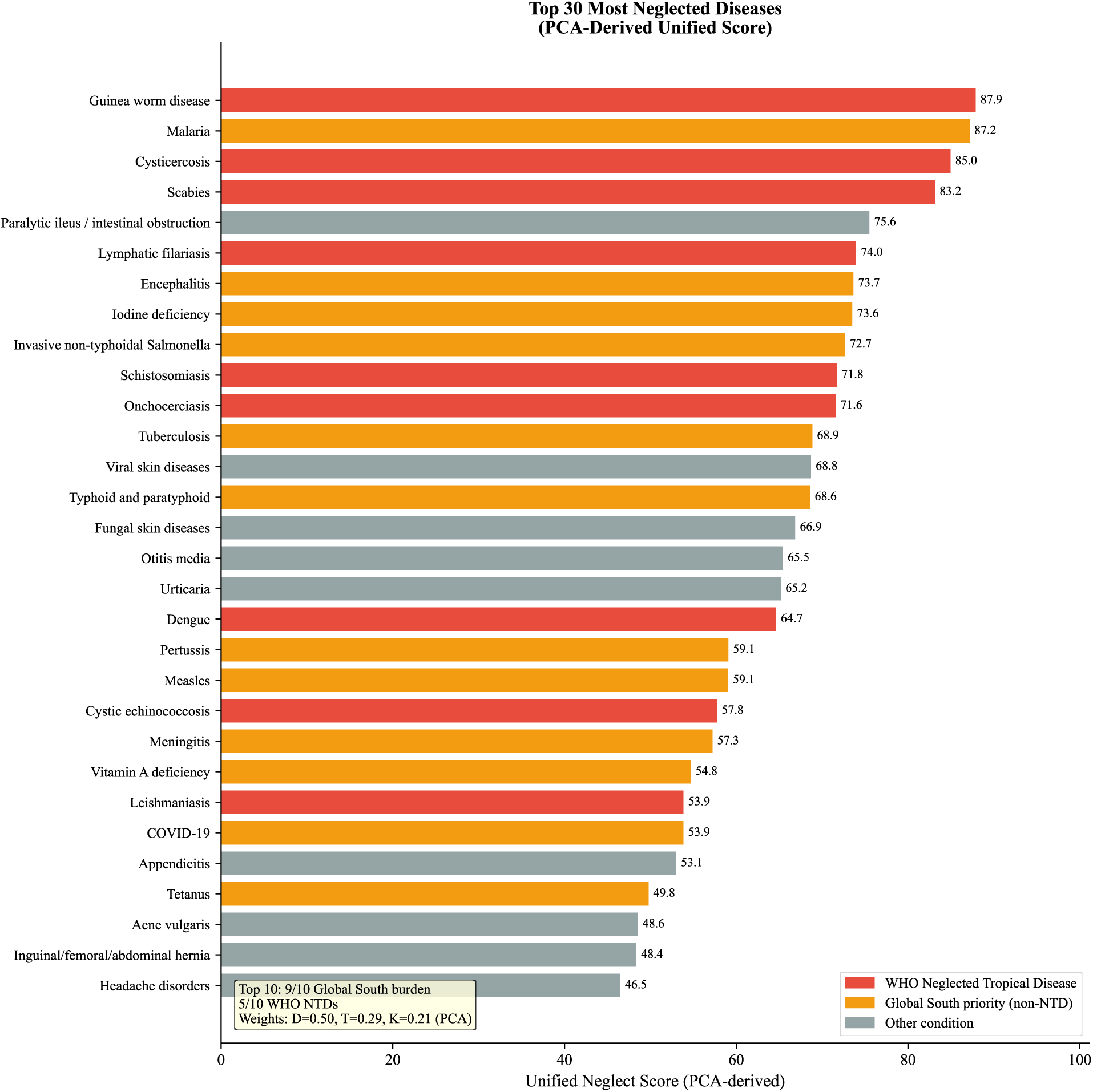
Nine of the ten most neglected diseases primarily burden the Global South. Top 30 diseases ranked by Unified Neglect Score. For the 86 diseases with complete data across all three dimensions, weights were derived from PCA (Discovery = 0.50, Translation = 0.29, Knowledge = 0.21; PC1 explains 63.3% of variance); for the remaining 89 diseases lacking clinical trial data, a two-dimension fallback was used (see Methods). Red bars indicate WHO Neglected Tropical Diseases (NTDs); orange bars indicate Global South priority diseases; grey bars indicate other conditions. Five of the top 10 are WHO-classified NTDs and nine have primary burden in the Global South.

The top ten most neglected diseases are further compared in **Table 1**. Nine have primary burden in the Global South, and five are classified as neglected tropical diseases by the WHO. The remainder, including malaria, encephalitis, iodine deficiency, and invasive non-typhoidal Salmonella, are likewise conditions of poverty and structural neglect. This convergence across independent dimensions (Discovery, Translation, and Knowledge) demonstrates cumulative neglect: these diseases are simultaneously absent from major biobanks, underrepresented in clinical trials, and semantically isolated in the research literature. As external validation, WHO NTD Roadmap diseases scored significantly higher on the Unified Neglect Score than non-NTD diseases (mean 49.4 vs. 27.6, P = 7.9 × 10⁻^4^, Mann-Whitney U), confirming that HEIM independently identifies internationally recognised priority diseases.

**Table 1.** Top Ten Most Neglected Diseases by Unified Score.

| Rank | Disease | WHO NTD | Unified Score | Gap Score | SII | Primary Burden Region |
| --- | --- | --- | --- | --- | --- | --- |
| 1 | Guinea worm disease | Yes | 87.9 | 95 | 0.00229 | Sub-Saharan Africa |
| 2 | Malaria | — | 87.2 | 95 | 0.00176 | Sub-Saharan Africa |
| 3 | Cysticercosis | Yes | 85.0 | 95 | 0.00203 | Latin America, Sub-Saharan Africa |
| 4 | Scabies | Yes | 83.2 | 95 | 0.00187 | Global South |
| 5 | Paralytic ileus and intestinal obstruction | — | 75.6 | 85 | 0.00193 | Global |
| 6 | Lymphatic filariasis | Yes | 74.0 | 95 | 0.00237 | Sub-Saharan Africa, South Asia |
| 7 | Encephalitis | — | 73.7 | 90 | 0.00140 | South Asia, Sub-Saharan Africa |
| 8 | Iodine deficiency | — | 73.6 | 82 | 0.00197 | South Asia, Sub-Saharan Africa |
| 9 | Invasive Non-typhoidal Salmonella (iNTS) | — | 72.7 | 80 | 0.00205 | Sub-Saharan Africa |
| 10 | Schistosomiasis | Yes | 71.8 | 95 | 0.00172 | Sub-Saharan Africa |

*Note: Five of ten are WHO-classified NTDs. Malaria, the second-highest scoring disease, is addressed separately by the WHO Global Malaria Programme. Seven of ten diseases lack sufficient clinical trial data; their Unified Score is computed from Discovery and Knowledge dimensions only (proportionally reweighted).*

By contrast, the diseases with lowest Unified Scores (greatest research equity) included breast cancer (0.5), colon and rectum cancer, and rheumatoid arthritis. These conditions share high publication volumes relative to disease burden and strong integration into the broader biomedical knowledge base. In particular, diseases with the highest absolute burden, such as ischemic heart disease (Unified Score 33.9, 188.4 million DALYs) and stroke (26.6, 160.5 million DALYs), received moderate Unified Scores reflecting that even well-researched conditions can show gaps when publication volume is assessed relative to their enormous disease burden.

### Temporal Trends: The Gap Is Not Closing

We analysed temporal evolution of research equity across the 26-year study period (2000-2025) using five-year rolling windows. Three findings emerged (**Extended Data Fig. 1**).

First, biobank research has become more concentrated rather than less. UK Biobank’s share of total biobank publications rose from 1% (2010–2014) to 57% (2020–2025), reflecting its scale and data accessibility after participant data became available. Global South biobanks’ collective share remained below 2% throughout.

Second, clinical trial site distribution showed no significant trend toward geographic equity. The HIC:LMIC site ratio fluctuated between 2.3 and 2.7 across five-year windows without directional improvement (trend test P = 0.41).

Third, neglected tropical diseases have not benefited from the expansion of biobank research. Despite a 47-fold increase in total biobank publications between 2000 and 2024, the share devoted to NTD-relevant diseases remained static at approximately 2% throughout the study period. In absolute terms, NTD publications in biobank research grew from fewer than 10 per year (2000-2005) to approximately 130 per year (2024), but this growth merely tracked the overall expansion of the field rather than reflecting any targeted increase in NTD research effort.

These trends demonstrate a sobering conclusion: despite decades of global health equity initiatives, the structural position of neglected diseases in the research landscape has not materially improved. The gap identified by HEIM is not a historical artefact but an ongoing feature of contemporary biomedical research. Incremental improvements in individual programmes have not translated into systemic change.

## Discussion

Neglected tropical diseases are not merely under-researched; they are structurally disconnected from the knowledge networks through which therapeutic innovation flows. Our semantic analysis of 13.1 million abstracts reveals that NTD research occupies isolated territory in biomedical knowledge space, 44% more distant from mainstream discourse than other conditions. This finding reframes research inequity: the problem is not only that onchocerciasis has averaged one clinical trial per year while breast cancer receives over 2,000, but that the science of neglected diseases is severed from the immunology, pharmacology, and genomics where breakthroughs originate. Our analysis suggests that the most neglected diseases are simultaneously absent from major biobanks, starved of clinical trials, and semantically disconnected from mainstream biomedicine. Each dimension reinforces the others: no biobank data means no drug targets; no trials means no approved therapies; no integration with mainstream science means no spillover from adjacent breakthroughs. For 1.5 billion people affected by the ten most neglected diseases, modern evidence-based medicine effectively does not exist.

The clinical consequences of this triple burden are measurable. The 2.4-fold gap in clinical trial intensity means that drug efficacy, optimal dosing regimens, and adverse effect profiles for Global South diseases rest on substantially thinner evidence bases. The dominance of observational designs (39.5% of NTD trials versus 23.2% for other conditions) means that even the limited evidence that exists is disproportionately descriptive rather than interventional; clinicians must extrapolate from studies with smaller sample sizes, shorter follow-up periods, and populations that may not reflect their patients. This evidence gap is compounded by the 2.5-fold concentration of trial sites in high-income countries: even when trials exist, they may underrepresent the genetic diversity, nutritional status, and co-infection patterns that influence treatment response in endemic regions.

The semantic isolation we identify poses particular risks as clinical medicine increasingly adopts AI-assisted tools for diagnosis and treatment selection ^48^. Clinical decision support systems, drug repurposing algorithms, and diagnostic AI are trained on literature corpora where neglected diseases are disconnected from the semantic networks of immunology, pharmacology, and genomics. When a clinician queries an AI system about potential drug interactions or differential diagnoses for a patient with lymphatic filariasis, the system draws on a knowledge base where that disease exists in isolation from mainstream therapeutic research. As these tools enter clinical practice, structural biases in the research literature risk becoming embedded in bedside decision-making, systematically disadvantaging patients with neglected conditions.

Beyond direct patient care, semantic isolation impedes therapeutic innovation. A disease that is disconnected from mainstream biomedical research cannot benefit from methodological advances in adjacent fields, i.e., novel drug delivery platforms, biomarker discovery techniques, or immunomodulatory strategies developed for conditions with greater research investment. The reverse is also true: by excluding diverse pathogen-host interactions from major biobanks, the biomedical community loses the biological insights that Global South diseases could contribute to understanding human immunity and drug response. This bidirectional isolation creates missed opportunities for drug repurposing: therapies effective against neglected diseases may go unrecognised because the conditions themselves are semantically invisible to researchers working in well-funded fields.

Our analysis of HIV/AIDS demonstrates that this pattern is reversible with sustained clinical and research investment. Over four decades, HIV research has become semantically integrated with virology, immunology, and pharmacology, with isolation decreasing by 24 per cent. Critically, this integration has translated into clinical advances: antiretroviral combinations that achieve durable viral suppression, pre-exposure prophylaxis that prevents transmission, and treatment protocols refined through large multinational trials. The mechanisms that achieved this involved dedicated funding streams such as PEPFAR and the Global Fund that mandated both clinical capacity building and cross-disciplinary research collaboration. Such mechanisms provide a template for addressing other neglected conditions. Semantic integration is not merely an academic metric; it correlates with the breadth and quality of therapeutic options available to clinicians.

To translate these findings into actionable intelligence, we developed an open-access interactive tool (https://manuelcorpas.github.io/17-EHR/) that goes beyond static visualisation. The tool provides three capabilities not available in existing equity frameworks. First, adjustable dimension weights allow users to modify the PCA-derived coefficients (Discovery, Translation, Knowledge) and observe how disease rankings respond in real time, enabling sensitivity exploration without rerunning the analysis. Second, a scenario builder models the effect of four policy interventions (e.g. increasing NTD research by 50%, equalising HIC/LMIC trial intensity, redistributing excess cancer trial capacity) on disease rankings, with Spearman rank correlations quantifying the magnitude of each shift. Third, a biobank comparison module allows side-by-side equity assessment of 2-5 IHCC biobanks by WHO region, identifying specific institutions where targeted investment would most reduce disease coverage gaps. These features collectively transform HEIM from a diagnostic framework into an instrument for research planning and accountability.

Addressing these inequities requires action at multiple levels. Funders should consider allocating a proportion of research portfolios to diseases with high Unified Neglect Scores, with requirements for the cross-disciplinary collaboration that reduces semantic isolation. Biobank consortia can adopt equity metrics to guide cohort recruitment toward populations that reflect global disease burden. Clinical trial sponsors should ensure that trials for Global South diseases include sufficient sites in endemic regions to capture relevant population characteristics. For clinicians, the HEIM dashboard (https://manuelcorpas.github.io/17-EHR/) provides a tool to identify which conditions face the greatest evidence gaps. The architecture of research inequity is now measurable and, as the HIV/AIDS trajectory demonstrates, it is reversible through deliberate investment in both clinical research capacity and knowledge integration.

Our analysis is subject to several limitations that warrant consideration. First, while the International Health Cohorts Consortium represents the largest coordinated biobank network globally, membership is voluntary. Consequently, non-member biobanks may differ systematically in their disease coverage, although our findings capture the most visible and well-resourced segment of the global infrastructure. Second, our reliance on PubMed inevitably favours English-language journals and may underrepresent research published in regional or non-English venues. However, this bias effectively strengthens our conclusion regarding semantic isolation within the dominant biomedical discourse that drives global funding and policy.

Third, clinical trial registration completeness varies by jurisdiction and has evolved over time, particularly following the 2005 ICMJE requirements ^49–51^. While this may affect absolute trial counts in the earlier years of our study period, the persistent geographic concentration we observe suggests a structural rather than administrative phenomenon. Fourth, our biobank analysis relies on indexed publication counts. This approach may underestimate research activity that does not result in indexed publications, yet publication remains the primary currency of scientific knowledge exchange and the mechanism by which biobanks influence clinical practice.

Finally, the Unified Neglect Score integrates three distinct dimensions using empirically derived PCA weights (Discovery = 0.50, Translation = 0.29, Knowledge = 0.21; PC1 explains 63.3% of variance). While alternative weightings could theoretically alter specific disease rankings, our sensitivity analyses demonstrate that the hierarchy of neglect is robust (**Extended Data Fig. 2**). Our semantic analysis relies on PubMedBERT, a single embedding model. While PubMedBERT was selected for its domain-specific pre-training on biomedical abstracts and established performance on biomedical NLP benchmarks, the narrow absolute range of SII values (0.0008 to 0.0033) means that alternative embedding models (e.g. SciBERT ^52^, BioLinkBERT ^53^) could in principle produce different absolute values. However, because our conclusions rest on relative rankings and group-level comparisons (NTD vs non-NTD) rather than absolute SII thresholds, and because the NTD isolation signal is large (Cohen’s d = 1.80), the core findings are unlikely to be model-dependent. Future work should confirm this with alternative embedding architectures. The clustering of Neglected Tropical Diseases at the bottom of the equity scale persists across a wide range of coefficient variations, confirming that our identification of compounding disadvantage is not an artefact of parameter selection.

The architecture of research neglect we have quantified is not inevitable. The HIV/AIDS trajectory proves that sustained, structured investment can integrate a disease into mainstream biomedicine within a generation. What HEIM provides is accountability. The neglect of Global South diseases can no longer be attributed to invisible market forces or unmeasurable complexity. It is now quantified, tracked, and attributable. Funders, institutions, and journals that claim commitment to global health equity now have a metric against which that commitment can be judged.

## Methods

### Study Design and Overview

We developed the Health Equity Informative Metrics (HEIM) framework to quantify research equity across three complementary dimensions: Discovery (biobank-derived research output), Translation (clinical trial activity), and Knowledge (semantic structure of the scientific literature). The framework was applied to 175 Level 3 disease categories from the Global Burden of Disease (GBD) 2021 taxonomy over the study period January 2000 to December 2025. By integrating bibliometric, clinical trial, and natural language processing analyses, HEIM captures the multi-stage nature of research neglect: from initial scientific investigation through translational activity to the structural organisation of accumulated knowledge. Each dimension produces disease-level and, where applicable, biobank-level scores that are normalised to a common 0-100 scale before integration into a Unified Neglect Score.

### Data Sources

Disease burden data were obtained from the Institute for Health Metrics and Evaluation (IHME) GBD 2021 study, comprising disability-adjusted life years (DALYs), deaths, and prevalence estimates for 175 Level 3 causes across 204 countries and territories. Thirty diseases were individually classified as Global South Priority conditions based on whether the disease disproportionately burdens low-and middle-income countries according to GBD 2021 DALY distributions, spanning infectious diseases, neglected tropical diseases, maternal and neonatal disorders, and nutritional deficiencies. Biobank metadata were obtained from the International Health Cohorts Consortium (IHCC) Global Cohort Atlas, yielding 70 biobanks with published research output across 29 countries. Biobank-linked publications were retrieved from PubMed/MEDLINE via the NCBI Entrez API (Biopython v1.79). For each of the 70 IHCC biobanks, we constructed Boolean queries combining 3-7 institutional name variants (e.g., (“UK Biobank”[All Fields] OR “United Kingdom Biobank”[All Fields] OR “U.K. Biobank”[All Fields])) to maximise recall. To circumvent PubMed’s 9,999-record return limit, queries were partitioned by publication date using [PDAT] field tags in year-by-year chunks (2000–2025), with automatic month-by-month subdivision for high-volume years. Records were fetched in XML format at a rate-limited 3 requests per second with retry logic (3 attempts, 5-second delay) for server errors. Title, abstract, journal, publication year, and MeSH descriptors were parsed for each record. Preprints and non-peer-reviewed records (medRxiv, bioRxiv, arXiv, Research Square, SSRN, and F1000Research) were excluded by journal-name string matching. Cross-biobank deduplication was performed on PubMed identifiers, with articles attributed to multiple biobanks retained as a single record. This yielded 38,595 unique publications. Each publication was mapped to 175 GBD 2021 disease categories via expert-curated MeSH descriptor vocabularies (e.g., Tuberculosis mapped to “Tuberculosis”, “Tuberculosis, Pulmonary”, “Mycobacterium tuberculosis”, “Tuberculosis, Multidrug-Resistant”, and “Latent Tuberculosis”), with substring matching (minimum 4 characters) as a fallback. All query logs, including exact search strings and per-biobank retrieval counts, are available in the project repository. Clinical trial records were obtained from the Aggregate Analysis of ClinicalTrials.gov (AACT) PostgreSQL database (aact-db.ctti-clinicaltrials.org). We queried five relational tables: studies (trial metadata, phase, status, enrolment, start and completion dates), conditions and browse_conditions (free-text and MeSH-mapped disease terms, respectively), countries (trial site locations), and sponsors (funding agency classification). Trials were included if their start date or first-posted date fell within 2000-2025. Records were fetched in chunks of 50,000 NCT identifiers to manage memory. Each trial was mapped to the 175 GBD 2021 Level 3 disease categories using a two-tier strategy: first, normalised MeSH-term matching against a curated dictionary of 189 disease-to-term correspondences (bidirectional substring comparison); second, keyword matching against free-text condition fields with word-boundary enforcement for short terms (≤4 characters). Disease mapping was parallelised across 24 CPU cores in batches of 50,000 trials. Facility records (n = 770,178 across 194 countries) were classified as high-income (HIC; 41 countries) or low-and middle-income (LMIC; 60 countries) using World Bank income groupings. Research intensity was computed as trials per million DALYs for each disease. This yielded 563,725 trials with at least one GBD disease mapping. For the Knowledge dimension, disease-specific PubMed abstracts were retrieved via the NCBI Entrez API using curated MeSH-based queries for each of the 175 GBD disease categories, partitioned by publication year (2000-2025) with a maximum of 100,000 PMIDs per disease per year. Abstracts were fetched in batches of 500 records in XML format; diseases were retained for semantic analysis if they met minimum quality thresholds (≥50 general PubMed abstracts, ≥70% year coverage across the 26-year window, and ≥95% abstract availability); all 175 GBD diseases met these criteria (the smallest corpus was Guinea worm disease with 429 abstracts). Note that these thresholds apply to the general PubMed literature for each disease, not to biobank-linked publications, which are counted separately in the Discovery dimension. This yielded 13,100,113 unique abstracts. Each abstract was encoded into a 768-dimensional vector using PubMedBERT (microsoft/BiomedNLP-PubMedBERT-base-uncased-abstract-fulltext) with mean pooling over non-padding tokens, tokenised to a maximum sequence length of 512, and processed in batches of 64 on Apple MPS (Metal Performance Shaders). Embeddings were stored in gzip-compressed HDF5 files with SHA-256 checksums for data integrity.

### Disease Mapping

Each data source was mapped to the GBD 2021 Level 3 taxonomy using a manually curated mapping of 175 GBD causes to MeSH vocabularies and keyword synonyms, embedded in the analysis pipeline. For biobank publications, MeSH terms from PubMed records were matched to GBD categories through exact and substring matching (minimum 4 characters). For clinical trials, a two-stage approach was used: MeSH terms assigned by ClinicalTrials.gov were matched first, followed by free-text condition names matched against curated keyword lists with word-boundary enforcement for short terms. Diseases were classified into GBD Level 2 groupings (e.g., neglected tropical diseases and malaria, cardiovascular diseases) and individually assigned a Global South priority flag based on whether the disease disproportionately burdens low-and middle-income countries (LMICs) according to GBD 2021 DALY distributions.

### Discovery Dimension Metrics

Three metrics characterise the Discovery dimension. The **Burden Score** provides a composite measure of disease severity: Burden Score = (0.5 × DALYs) + (50 × Deaths) + [10 × log₁₀(Prevalence)] where DALYs and Deaths are expressed in millions and Prevalence in millions of cases. The three components span markedly different scales across the 175 GBD diseases: DALYs range from <0.01 to 212.0 million, deaths from 0 to 9.0 million, and prevalence from near-zero to 3,692 million. Coefficients were therefore selected to normalise each component to a comparable numerical range so that no single dimension dominates the composite. Multiplying DALYs by 0.5 yields a contribution range of 0-106; multiplying deaths by 50 yields 0-450; and applying log₁₀ to prevalence (compressing a 3,700-fold spread to a ∼6-unit range) and multiplying by 10 yields approximately-46 to 36. Deaths thus receive the largest effective weight (∼50.7% of the total score on average), reflecting the prioritisation of lethality as an equity indicator; DALYs contribute ∼21.8% and prevalence ∼27.5%. The log-transformation of prevalence prevents conditions with very high case counts but low per-case severity (e.g., oral disorders, headache disorders) from dominating the composite. Resulting rankings were verified for validity against GBD 2021 priorities: high-burden conditions such as ischaemic heart disease, COVID-19, and neonatal disorders ranked at the top, whilst lower-burden conditions such as anxiety disorders and bipolar disorder ranked at the bottom. Sensitivity analysis varying each coefficient independently by ±20% and ±50% confirmed complete rank stability (Spearman ρ > 0.999 under all 12 perturbation schemes; **Extended Data Fig. 2**). This insensitivity arises because the rank order is dominated by large inter-disease differences in DALYs and deaths, which span four orders of magnitude and are preserved regardless of precise coefficient values. When only DALYs were available, a log-scaled fallback (10 × log₁₀(DALYs + 1)) was used to ensure comparability. The **Research Gap Score** (0-100) employs a three-tier system: (i) a zero-publication penalty (Gap = 95) for diseases with no biobank output, reflecting complete research absence; (ii) category-specific thresholds with stricter criteria for infectious and neglected tropical diseases, which require higher absolute publication volumes given their global burden (for example, malaria and tuberculosis each require at least 50 publications to avoid threshold-based elevation); and (iii) burden-normalised research intensity (publications per million DALYs) for remaining conditions, scored relative to the best-performing disease in the dataset. An additional +10 penalty is applied to Global South priority diseases with fewer than 50 publications. Gap severity categories are Critical (>70), High (50-70), Moderate (30-50), and Low (<30). These thresholds were anchored to empirically interpretable benchmarks: Critical (>70) corresponds to diseases with fewer than five publications across all 70 biobanks, representing near-complete research absence; High (50-70) captures diseases with minimal but non-zero coverage; Moderate (30-50) marks the transition to diseases receiving substantive, if incomplete, attention; and Low (<30) identifies diseases with at least moderate research representation. Alternative cut-point schemes (±10 points) were tested across 20 threshold-penalty combinations; the underlying Gap Score rankings remained highly stable (Spearman ρ > 0.9998), with the proportion of diseases classified as Critical or High varying by fewer than 4 percentage points. The **Equity Alignment Score** (EAS) integrates biobank-level performance: EAS = 100 − (0.4 × Gap_Severity + 0.3 × Burden_Miss + 0.3 × Capacity_Penalty) where Gap_Severity is a weighted count of gap categories (Critical = 4, High = 2, Moderate = 1), Burden_Miss is the proportion of DALYs in diseases with ≤2 publications, and Capacity_Penalty is the inverse of per-disease publication coverage. Gap_Severity was assigned the largest weight (0.4) as the most direct measure of research neglect; the remaining weight was split equally (0.3 each) between Burden_Miss, which captures whether high-burden diseases lack coverage, and Capacity_Penalty, which captures breadth of the disease portfolio. Sensitivity analysis across seven alternative weight schemes, ranging from equal weighting (0.33/0.33/0.34) to single-component dominance (0.6/0.2/0.2), yielded Spearman ρ ≥ 0.991 for all pairwise rank comparisons, confirming that biobank rankings are robust to weight reallocation. EAS ranges from 0 to 100, with higher scores indicating better alignment. Category thresholds were derived from the observed EAS distribution across 70 IHCC biobanks (median = 28.4, interquartile range 18.7-37.1): the High threshold of 70 was set at approximately 2 standard deviations above the mean, ensuring that only biobanks with demonstrably comprehensive disease coverage qualified; the Moderate threshold of 40 corresponds approximately to the 75th percentile, distinguishing above-average from below-average performers. Shifting both thresholds by ±5 points altered the number of biobanks in each category by at most 3, and the sole biobank classified as High (UK Biobank, EAS = 84.6) remained unchanged across all threshold schemes tested. EAS categories are High (≥70), Moderate (40-69), and Low (<40).

### Translation Dimension Metrics

Clinical trial equity was assessed through one scoring metric and two characterisation analyses. The scoring metric, **research intensity**, was calculated as the number of registered trials per million DALYs for each disease category, enabling direct comparison of translational investment relative to disease burden. DALYs were chosen as the denominator because they integrate both mortality and morbidity into a single measure of total health loss, permitting comparison across diseases with different fatality profiles. For the **Unified Score**, research intensity was inverted (1/(intensity + 1)) so that diseases with fewer trials per DALY received higher neglect scores, then min–max normalised to [0, 1] across all 175 diseases. Two additional analyses contextualise the scoring metric. **Geographic distribution** was quantified by classifying each of 770,178 trial facility records as high-income country (HIC) or LMIC based on World Bank income classifications, yielding an HIC:LMIC site ratio. Temporal trends were analysed using non-overlapping five-year periods (2000-2005, 2006-2010, 2011-2015, 2016-2020, 2021-2025) to assess whether the share of trials addressing Global South priority diseases has changed over the study period. A total of 563,725 trial records from ClinicalTrials.gov (accessed via the AACT database) covering 194 countries were included; 89 disease categories had at least one mapped trial and were retained for Translation dimension analysis.

### Knowledge Dimension Metrics

Semantic analysis ^54^ used PubMedBERT (microsoft/BiomedNLP-PubMedBERT-base-uncased-abstract-fulltext, version 1.1) to generate 768-dimensional vector representations for each abstract, with a maximum token length of 512 and batch size of 64. Disease-level centroids were computed as the mean embedding across all abstracts for each condition. The primary scoring metric is the **Semantic Isolation Index (SII)**, which measures the mean cosine distance from each disease’s centroid to the centroids of its k = 100 nearest-neighbour diseases in the embedding space; higher values indicate research that is more disconnected from mainstream biomedical literature. We set k = 100 (57% of the 175 diseases) to capture broad-spectrum isolation rather than local neighbourhood effects; because the embedding space is high-dimensional (768 dimensions) and disease centroids cluster tightly, a large k is needed to resolve meaningful differences in isolation. The resulting SII values range from 0.0008 to 0.0033 across diseases, a narrow absolute range reflecting the high overall similarity of biomedical language, but one that nonetheless discriminates between well-integrated diseases (e.g., cardiovascular conditions, SII ≈ 0.0008) and isolated ones (e.g., neglected tropical diseases, SII ≈ 0.0025–0.0033). For the Unified Score, SII was min-max normalised to [0, 1] across all 175 diseases. Three complementary characterisation metrics were also computed but were not included in the Unified Score, to maintain interpretive simplicity. **Knowledge Transfer Potential (KTP)** captures cross-disease similarity as the mean cosine similarity between a disease’s centroid and the centroids of its top 10% (n ≈ 17) most similar conditions, reflecting the potential for methodological and conceptual spillover. The **Research Clustering Coefficient (RCC)** quantifies within-disease embedding variance as the mean cosine distance from individual abstracts to their disease centroid, with higher values indicating a more dispersed research agenda. Temporal semantic drift was measured as the cosine distance between consecutive yearly centroids for each disease, capturing how research focus evolves over time.

### Unified Neglect Score

To identify diseases experiencing compounding disadvantage across all three dimensions, we computed a Unified Neglect Score integrating normalised metrics from each dimension: Unified Score = (0.50 × Discovery) + (0.29 × Translation) + (0.21 × Knowledge) Each dimension score was normalised to a 0-100 scale before integration: Discovery scores used the Gap Score, Translation scores used the inverse of research intensity (scaled to 0-100), and Knowledge scores used the Semantic Isolation Index (scaled to 0-100). Dimension weights were derived empirically using principal component analysis (PCA) on the 86 diseases with data across all three dimensions. The first principal component explained 63.3% of total variance, with normalised loadings of 0.50 (Discovery), 0.29 (Translation), and 0.21 (Knowledge), indicating that Discovery-stage neglect contributes most to overall neglect variance. For 89 diseases lacking sufficient clinical trial data (Translation dimension), the Unified Score was computed from Discovery and Knowledge dimensions only, with weights proportionally rescaled to 0.71 (Discovery) and 0.29 (Knowledge). Sensitivity analyses examined alternative weighting schemes and confirmed that NTD rankings were stable (Spearman’s ρ > 0.975 across all schemes; **Extended Data Fig. 2**). The maximum Unified Score observed was 87.9 (for Guinea worm disease). No disease simultaneously received the highest possible scores on all three dimensions, reflecting the partial independence of the three dimensions.

### Statistical Analysis

Group comparisons (e.g., NTD vs. non-NTD, Global South priority vs. non-priority) used Welch’s t-test with Cohen’s d effect sizes. Correlations between metrics (e.g., Gap Score vs. Semantic Isolation Index) were assessed using Pearson’s r. Multiple comparisons were corrected using the Benjamini-Hochberg procedure with a false discovery rate threshold of q < 0.05. Sensitivity analyses were conducted by comparing PCA-derived weights against equal weighting and 1,000 random Dirichlet-distributed weight vectors; rank-order stability was assessed using Spearman’s ρ (all ρ > 0.975). Dimensionality reduction for semantic visualisation used Uniform Manifold Approximation and Projection (UMAP) with n_neighbours = 15, min_dist = 0.1, and cosine distance metric.

### External Validation

To assess whether HEIM independently identifies internationally recognised priority diseases, we compared Unified Neglect Scores against the 20 diseases prioritised by the WHO Neglected Tropical Diseases Roadmap 2021-2030 ^55^: Buruli ulcer, Chagas disease, dengue, dracunculiasis (Guinea worm disease), echinococcosis, foodborne trematodiases, human African trypanosomiasis, leishmaniasis, leprosy, lymphatic filariasis, mycetoma, onchocerciasis, rabies, scabies, schistosomiasis, soil-transmitted helminthiases (ascariasis, hookworm disease, trichuriasis), snakebite envenoming, taeniasis/cysticercosis, trachoma, and yaws. Of these 20, 17 mapped to individual GBD Level 3 categories in our dataset. Echinococcosis mapped as cystic echinococcosis; ascariasis, hookworm disease, and trichuriasis lacked individual GBD Level 3 entries and were excluded. We tested whether WHO NTDs ranked significantly higher on the Unified Neglect Score than non-NTD diseases using a one-sided Mann-Whitney U test.

## Code and Data Availability

All analysis code, including data retrieval, disease mapping, metric computation, embedding generation, and figure generation pipelines, is available at https://github.com/manuelcorpas/17-EHR. **An interactive dashboard for exploring HEIM metrics across all three dimensions is available at** https://manuelcorpas.github.io/17-EHR/. Tool capabilities are described in the Discussion. GBD 2021 data are publicly available from IHME (https://ghdx.healthdata.org/). Clinical trial data are available from the AACT database (https://aact.ctti-clinicaltrials.org/). IHCC biobank registry data are available from the Global Cohort Atlas.

## Data Availability

All data used in this study are derived from publicly available sources. Global Burden of Disease 2021 data are available from the Institute for Health Metrics and Evaluation (IHME). Clinical trial data were obtained from the AACT ClinicalTrials.gov database. Bibliometric data were retrieved from PubMed/MEDLINE. All analysis code, derived metrics, and visualisation pipelines are openly available.

https://github.com/manuelcorpas/17-EHR

https://manuelcorpas.github.io/17-EHR/

https://ghdx.healthdata.org/

https://aact.ctti-clinicaltrials.org/

## Acknowledgements

We thank the International Health Cohorts Consortium for providing biobank registry data, the Clinical Trials Transformation Initiative for maintaining the AACT database, and the National Library of Medicine for PubMed access.

## Author Contributions

**M.C.:** Conceptualisation, Methodology, Software, Formal Analysis, Data Curation, Writing (Original Draft, Visualisation, Project Administration). **M.F.:** Methodology, Validation, Writing, Review & Editing. **J.V.-S.:** Writing, Review & Editing. **S.B.:** Writing, Review & Editing. **S.F.:** Conceptualisation, Validation, Writing, Review & Editing. **H.G.:** Writing, Review & Editing.

## Additional Information

Correspondence and requests for materials should be addressed to M.C.

## Extended Data Figures

**Extended Data Fig. 1.**
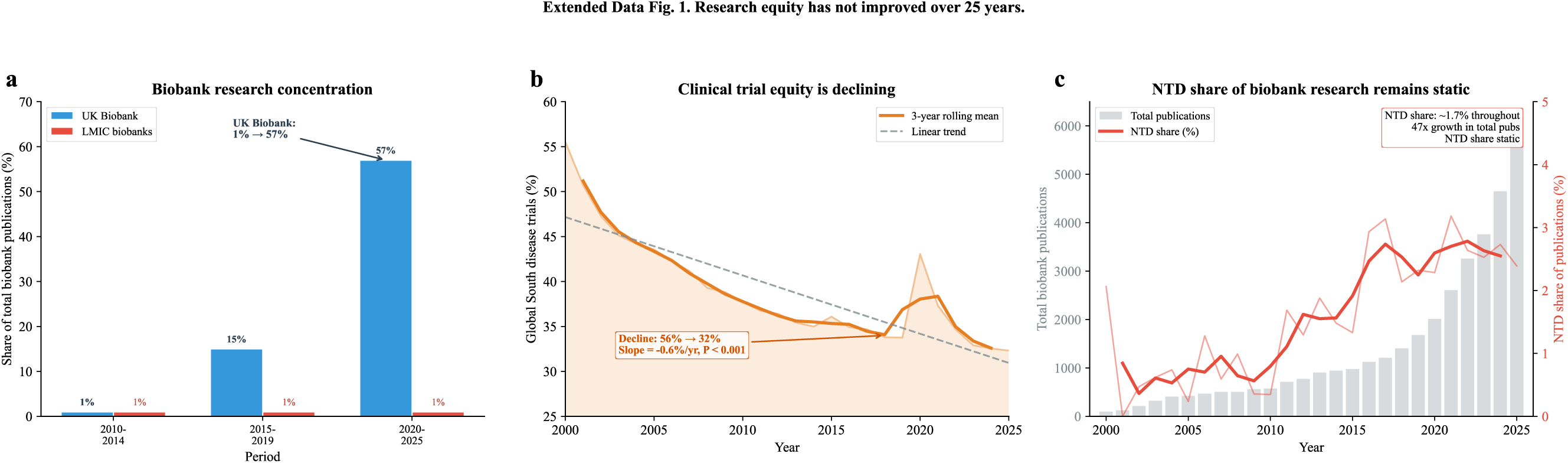
Research equity has not improved over 25 years. (a) UK Biobank’s share of total biobank publications rose from 1% (2010-2014) to 57% (2020-2025) after participant data became available, while LMIC biobanks remained below 2% throughout. (b) The share of clinical trials addressing Global South priority diseases declined from 56% (2000) to 32% (2025), with a linear trend of −0.6 percentage points per year (P < 0.001). (c) Despite a 47-fold increase in total biobank publications, the proportion addressing neglected tropical diseases remained static at approximately 2%.

**Extended Data Fig. 2.**
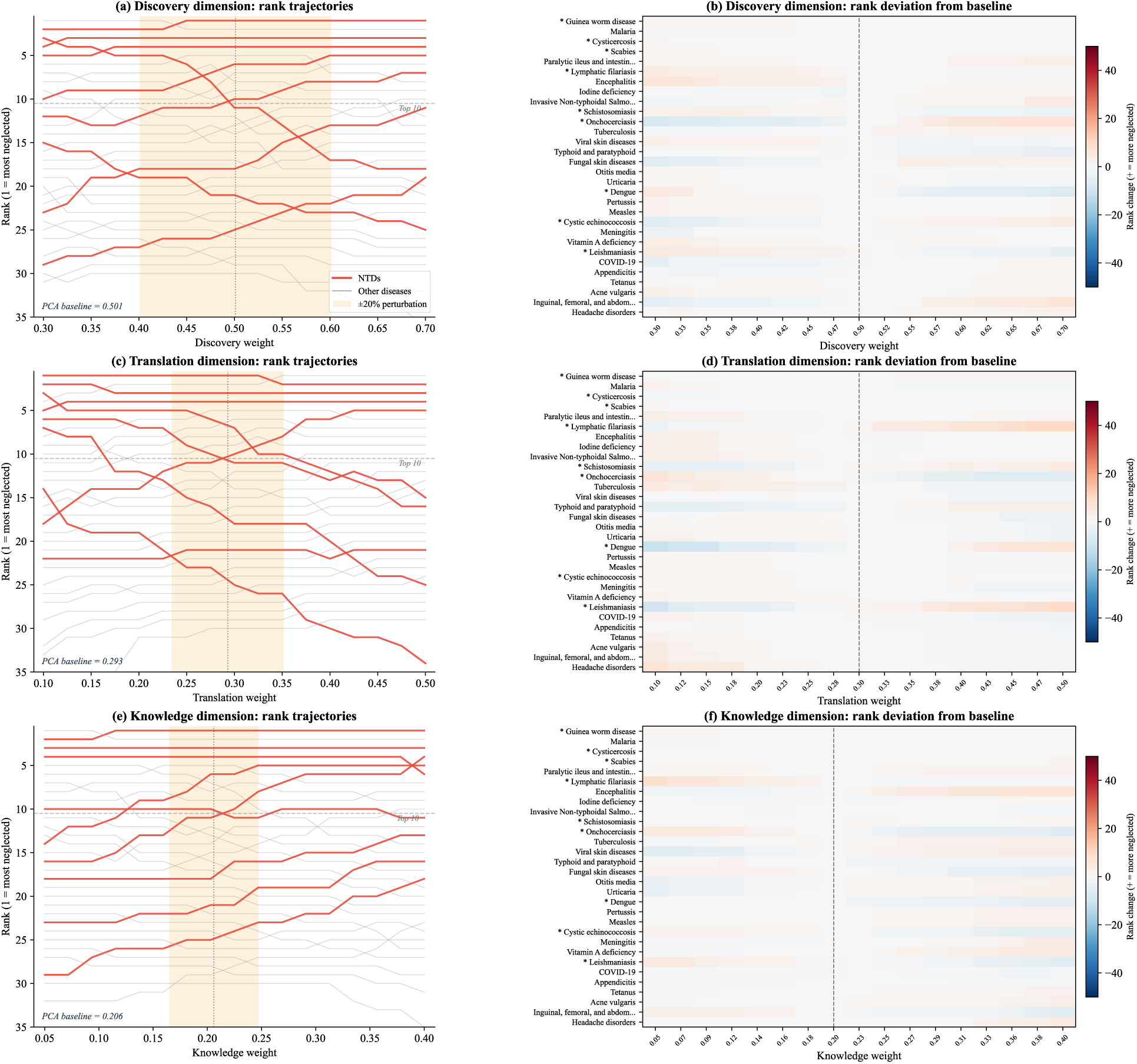
Disease neglect rankings are robust to alternative weighting schemes. Each row independently varies one dimension’s weight across 17 schemes while keeping the other two in their PCA-derived ratio; the left column shows rank trajectories for the 30 most neglected diseases and the right column shows rank deviations from the PCA baseline (Discovery = 0.50, Translation = 0.29, Knowledge = 0.21). (a, b) Discovery weight varied from 0.30 to 0.70 (Spearman’s rho = 0.976–1.000). (c, d) Translation weight varied from 0.10 to 0.50 (rho = 0.992–1.000); 2D diseases lacking clinical-trial data are unaffected, explaining the higher stability. (e, f) Knowledge weight varied from 0.05 to 0.40 (rho = 0.975–1.000). Red lines and asterisks indicate NTDs; grey lines indicate other diseases. The orange shaded region marks the +/-20% perturbation range around each baseline weight. In the heatmaps, red indicates worsened rank (more neglected) and blue indicates improved rank (less neglected); the dashed line marks the baseline column. Rankings remain highly stable across all three dimensions, with NTDs consistently occupying the highest neglect positions and most deviations negligible for the top 10 diseases (minimum rho = 0.975 across 51 total schemes).

**Extended Data Fig. 3.**
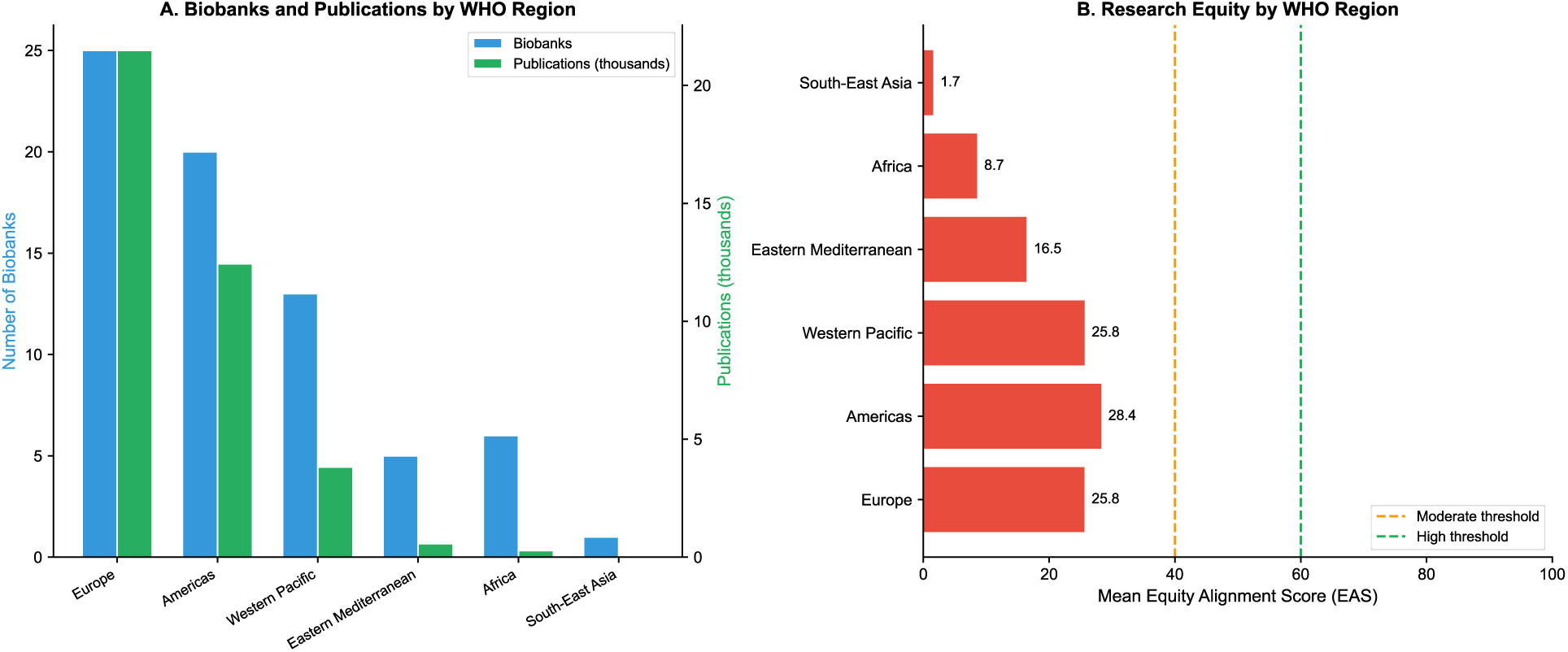
Research output concentrates in Europe and the Americas. (A) Biobanks and publications by WHO region. (B) Research equity by WHO region showing mean Equity Alignment Scores.

## References

1. de Almeida, A. B. & Freedman, D. O. Epidemiology and immunopathology of bancroftian filariasis. Microbes and Infection 1, 1015–1022 (1999).

2. Connolly, J. J. et al. The International Health Cohorts Consortium (IHCC) advances population health research and genomic discovery. Commun Med 5, 366 (2025).

3. Ndubuisi, N. E. Noncommunicable Diseases Prevention In Low-and Middle-Income Countries: An Overview of Health in All Policies (HiAP). Inquiry 58, 0046958020927885 (2021).

4. Sirugo, G., Williams, S. M. & Tishkoff, S. A. The Missing Diversity in Human Genetic Studies. Cell 177, 26–31 (2019).

5. Schmallenbach, L. et al. Global distribution of research efforts, disease burden, and impact of US public funding withdrawal. Nat Med 31, 3101–3109 (2025).

6. Arhel, N. J. Fostering better science by releasing biomedical research and innovation from the grip of rich nations. PLOS Glob Public Health 5, e0004709 (2025).

7. Hotez, P. J. The poverty-related neglected diseases: Why basic research matters. PLOS Biology 15, e2004186 (2017).

8. Viergever, R. F. The mismatch between the health research and development (R&D) that is needed and the R&D that is undertaken: an overview of the problem, the causes, and solutions. Glob Health Action 6, 10.3402/gha.v6i0.22450 (2013).

9. Grijsen, M. L., et al. Rethinking neglected tropical diseases: A shift towards more inclusive and equitable terminology. PLOS Glob Public Health 5, e0004094 (2025).

10. Zimmerman, A. et al. Investing in a global pooled-funding mechanism for late-stage clinical trials of poverty-related and neglected diseases: an economic evaluation. BMJ Glob Health 8, e011842 (2023).

11. Bleich, S. N., Jarlenski, M. P., Bell, C. N. & LaVeist, T. A. Health Inequalities: Trends, Progress, and Policy. Annu Rev Public Health 33, 7–40 (2012).

12. National Academies of Sciences, E. et al. The Root Causes of Health Inequity. In Communities in Action: Pathways to Health Equity (National Academies Press (US), 2017).

13. Rad, J. Health inequities: a persistent global challenge from past to future. Int J Equity Health 24, 148 (2025).

14. Kumar, R., Khosla, R. & McCoy, D. Decolonising global health research: Shifting power for transformative change. PLOS Glob Public Health 4, e0003141 (2024).

15. Hardeman, R. R., Homan, P. A., Chantarat, T., Davis, B. A. & Brown, T. H. Improving The Measurement Of Structural Racism To Achieve Antiracist Health Policy. Health Aff (Millwood) 41, 179–186 (2022).

16. Bekker, L.-G. et al. Advancing global health and strengthening the HIV response in the era of the Sustainable Development Goals: the International AIDS Society—Lancet Commission. The Lancet 392, 312–358 (2018).

17. Elendu, C. et al. Shaping sustainable paths for HIV/AIDS funding: a review and reminder. Ann Med Surg (Lond) 87, 1415–1445 (2025).

18. Fajardo-Ortiz, D. et al. The emergence and evolution of the research fronts in HIV/AIDS research. PLoS One 12, e0178293 (2017).

19. Scherer, E., Douek, D. & McMichael, A. 25 years of HIV research on virology, virus restriction, immunopathogenesis, genes and vaccines. Clin Exp Immunol 154, 6–14 (2008).

20. Donkin, A., Goldblatt, P., Allen, J., Nathanson, V. & Marmot, M. Global action on the social determinants of health. BMJ Glob Health 3, (2018).

21. Graves, J. L., Kearney, M., Barabino, G. & Malcom, S. Inequality in science and the case for a new agenda. Proc Natl Acad Sci U S A 119, e2117831119 (2022).

22. Petersen, O. H. Inequality of Research Funding between Different Countries and Regions is a Serious Problem for Global Science. Function (Oxf) 2, zqab060 (2021).

23. Dorantes-Gilardi, R. et al. Quantifying the impact of biobanks and cohort studies. Proc Natl Acad Sci U S A 122, e2427157122.

24. Bahcall, O. G. UK Biobank — a new era in genomic medicine. Nat Rev Genet 19, 737–737 (2018).

25. Khodursky, S., Mimouni, N. & Levin, M. G. Recent developments in population biobanks and the genetic architecture of complex disease. Hum Mol Genet ddaf036 (2025) doi:10.1093/hmg/ddaf036.

26. Fatumo, S. et al. A roadmap to increase diversity in genomic studies. Nat. Med. 28, 243–250 (2022).

27. Corpas, M. et al. Bridging genomics’ greatest challenge: The diversity gap. Cell Genom 5, 100724 (2025).

28. Popejoy, A. B. & Fullerton, S. M. Genomics is failing on diversity. Nature 538, 161–164 (2016).

29. Smyth, B. et al. Inequities in the global representation of sites participating in large, multicentre dialysis trials: a systematic review. BMJ Glob Health 4, e001940 (2019).

30. Marshall, I. J. et al. State of the evidence: a survey of global disparities in clinical trials. BMJ Glob Health 6, e004145 (2021).

31. Atal, I., Trinquart, L., Ravaud, P. & Porcher, R. A mapping of 115,000 randomized trials revealed a mismatch between research effort and health needs in non–high-income regions. Journal of Clinical Epidemiology 98, 123–132 (2018).

32. Sweileh, W. M. A bibliometric analysis of global research output on health and human rights (1900–2017). Glob Health Res Policy 3, 30 (2018).

33. Ganti, L., Persaud, N. A. & Stead, T. S. Bibliometric analysis methods for the medical literature. Academic Medicine & Surgery (2025) doi:10.62186/001c.129134.

34. Evans, J. A., Shim, J.-M. & Ioannidis, J. P. A. Attention to Local Health Burden and the Global Disparity of Health Research. PLoS One 9, e90147 (2014).

35. Stuckler, D., King, L., Robinson, H. & McKee, M. WHO’s budgetary allocations and burden of disease: a comparative analysis. Lancet 372, 1563–1569 (2008).

36. Nwaka, S. et al. Advancing Drug Innovation for Neglected Diseases—Criteria for Lead Progression. PLoS Negl Trop Dis 3, e440 (2009).

37. Calderón, F., Fairlamb, A. H., Strange, M., Williams, P. & Nathan, C. F. Surmounting structural barriers to tackle endemic infectious diseases. J Exp Med 218, e20211418 (2021).

38. Ong, M. A comprehensive framework identifying barriers to global health R&D innovation and access. BMJ Glob Health 8, (2023).

39. Trends and Charts on Registered Studies | ClinicalTrials.gov. https://clinicaltrials.gov/about-site/trends-charts.

40. Institute For Health Metrics And Evaluation. Global Burden of Disease Study 2021 (GBD 2021) Cause, REI, and Location Hierarchies. Institute for Health Metrics and Evaluation 10.6069/G44H-ZC04 (2024).

41. Colditz, G. A., Manson, J. E. & Hankinson, S. E. The Nurses’ Health Study: 20-Year Contribution to the Understanding of Health Among Women. Journal of Women’s Health 6, 49–62 (1997).

42. Writing Group for the Women’s Health Initiative Investigators. Risks and Benefits of Estrogen Plus Progestin in Healthy Postmenopausal WomenPrincipal Results From the Women’s Health Initiative Randomized Controlled Trial. JAMA 288, 321–333 (2002).

43. Milani, L. et al. The Estonian Biobank’s journey from biobanking to personalized medicine. Nat Commun 16, 3270 (2025).

44. Okpechi, I. G. et al. Task shifting roles, interventions and outcomes for kidney and cardiovascular health service delivery among African populations: a scoping review. BMC Health Serv Res 23, 446 (2023).

45. Praziquantel for Schistosomiasis: Single-Drug Metabolism Revisited, Mode of Action, and Resistance. Antimicrobial Agents and Chemotherapy https://journals.asm.org/doi/10.1128/aac.02582-16.

46. Gu, Y. et al. Domain-Specific Language Model Pretraining for Biomedical Natural Language Processing. ACM Trans. Comput. Healthcare 3, 1–23 (2022).

47. Adams, J. & Light, R. Mapping Interdisciplinary Fields: Efficiencies, Gaps and Redundancies in HIV/AIDS Research. PLoS One 9, e115092 (2014).

48. Ouanes, K. & Farhah, N. Effectiveness of Artificial Intelligence (AI) in Clinical Decision Support Systems and Care Delivery. J Med Syst 48, 74 (2024).

49. Gresham, G., Meinert, J. L., Gresham, A. G., Piantadosi, S. & Meinert, C. L. Update on the clinical trial landscape: analysis of ClinicalTrials.gov registration data, 2000–2020. Trials 23, 858 (2022).

50. Moja, L. P. et al. Compliance of clinical trial registries with the World Health Organization minimum data set: a survey. Trials 10, 56 (2009).

51. Al-Durra, M., Nolan, R. P., Seto, E. & Cafazzo, J. A. Prospective registration and reporting of trial number in randomised clinical trials: global cross sectional study of the adoption of ICMJE and Declaration of Helsinki recommendations. BMJ 369, m982 (2020).

52. Beltagy, I., Lo, K. & Cohan, A. SciBERT: A Pretrained Language Model for Scientific Text. in Proceedings of the 2019 Conference on Empirical Methods in Natural Language Processing and the 9th International Joint Conference on Natural Language Processing (EMNLP-IJCNLP) (eds Inui, K., Jiang, J., Ng, V. & Wan, X.) 3615–3620 (Association for Computational Linguistics, Hong Kong, China, 2019). doi:10.18653/v1/D19-1371.

53. Yasunaga, M., Leskovec, J. & Liang, P. LinkBERT: Pretraining Language Models with Document Links. Preprint at 10.48550/arXiv.2203.15827 (2022).

54. Khakimova, A., Rahim, F. & Zolotarev, O. Bibliometric and Semantic Analysis of the Global Research on Biomarkers in Personalized Medicine. doi:10.2174/18753183-v12-e220926-2022-3.

55. Malecela, M. N. & Ducker, C. A road map for neglected tropical diseases 2021–2030. Trans R Soc Trop Med Hyg 115, 121–123 (2021).

